# Development and Multinational Validation of Artificial Intelligence-Enabled ASCVD Risk Stratification Using Electrocardiograms

**DOI:** 10.64898/2026.01.05.26343465

**Authors:** Bruno Batinica, Evangelos K Oikonomou, Aline F Pedroso, Arya Aminorroaya, Dhruva Biswas, Sandhi M Barreto, Luisa C C Brant, Antonio L P Ribeiro, Lovedeep S Dhingra, Rohan Khera

**Affiliations:** Department of Internal Medicine, Section of Cardiovascular Medicine, Yale School of Medicine, New Haven, CT 06510, USA; Cardiovascular Data Science (CarDS) Lab, Yale School of Medicine, New Haven, CT 06510, USA; Department of Preventive Medicine, School of Medicine, Faculdade de Medicina, Universidade Federal de Minas Gerais, Belo Horizonte, Brazil; Department of Internal Medicine, Faculdade de Medicina, Universidade Federal de Minas Gerais, Belo Horizonte, Brazil; Telehealth Center and Cardiology Service, Hospital das Clínicas, Universidade Federal de Minas Gerais, Belo Horizonte, Brazil; Center for Outcomes Research and Evaluation (CORE), Yale New Haven Hospital, New Haven, CT 06510, USA; Section of Biomedical Informatics and Data Science, Yale School of Medicine, New Haven, CT 06510, USA; Section of Health Informatics, Department of Biostatistics, Yale School of Public Health, New Haven, CT 06510, USA

**Keywords:** Atherosclerotic cardiovascular disease, risk prediction, artificial intelligence, electrocardiogram

## Abstract

**Aims:** Despite the availability of clinical risk scores for atherosclerotic cardiovascular disease (ASCVD), their use is limited because the required predictor data are often missing. We developed and validated ECG-ASCVD, a scalable risk prediction paradigm that utilizes ECGs to target ASCVD risk factor assessment.

**Methods:** Adults aged 30-79 who had undergone a clinical ECG were identified in the Yale New Haven Health System (YNNHS) and a state death index. We developed ECG-ASCVD-12, ECG-ASCVD-IMAGE, and ECG-ASCVD-1 to predict time-to-ASCVD from 12-lead ECG signals, ECG images, and lead-1 signals, respectively. Model performance was assessed in held-out individuals without prior ASCVD and in two external prospective cohorts, ELSA-Brasil (ELSA) and the UK Biobank (UKB). We then simulated the deployment of ECG-ASCVD in a random sample of 100,000 adults at YNHHS.

**Results:** The development cohort included 363,788 individuals (median age, 57.1 [45.5-67.2] years; 48% Women). The YNHHS, ELSA, and UKB test cohorts included 83,917, 10,934, and 54,166 individuals, respectively. ECG-ASCVD-12 demonstrated generalizable discrimination (C-index: 0.684 to 0.746) and remained independently associated with ASCVD (adjusted hazard ratio: 1.23-1.34 per SD) after adjusting for PREVENT scores (C-index: 0.696-0.782) across the validation cohorts. ECG-ASCVD-IMAGE performed similarly (C-index: 0.673-0.748) while ECG-ASCVD-1 had modestly lower performance (C-index: 0.671-0.735). Simulated deployment suggested that ECG-ASCVD could enable the detection of high ASCVD risk patients who lack the data required for PREVENT.

**Conclusion:** We developed an ECG-ASCVD toolkit and validated it across diverse multinational cohorts. These results highlight the potential utility of resting ECG information for predicting ASCVD risk, enabling targeted screening.

## Introduction

Although cardiovascular risk prediction models have revolutionized the primary prevention of ASCVD, in practice, many individuals never undergo formal risk assessment.^1–3^ Current clinical risk models require comprehensive risk factor information, including clinical evaluation, detailed medical history, and laboratory tests for score calculation, which limits their utilization.^4^ Individuals who are not risk-assessed are less likely to receive appropriate therapy, and most who ultimately experience an ASCVD event are not prescribed or taking guideline-recommended treatment.^5,6^ As ASCVD remains the leading cause of death and disability worldwide, there is a critical need for accurate, accessible, and scalable approaches to help target ASCVD risk stratification.^7^

A promising tool to help improve ASCVD risk prediction is the utilization of artificial intelligence (AI) applied to electrocardiograms (ECGs), which are low-cost, non-invasive, and widely available tests already integrated into routine clinical care. ECGs are near ubiquitous in clinical practice for the workup of individuals with presumed cardiovascular disease, with over 300 million studies performed each year.^8^ There has been long-standing recognition that parameters derived from routine 12-lead ECG interpretation contain information that can predict future cardiovascular disease.^9,10^ Recent AI advances have enabled the extraction of even richer latent prognostic information from ECGs for several cardiovascular diseases.^11–15^ ECG could be a single test to guide the more comprehensive assessment needed for risk prediction.

While prior studies have demonstrated that predictive information regarding ASCVD can be extracted from 12-lead ECG voltage data, how this information compares with contemporary clinical ASCVD risk models (such as Predicting Risk of Cardiovascular Disease EVENTs (PREVENT)), or how models developed for more accessible ECG modalities (ECG images and single-lead signals) perform remains unexplored.^16–18^ In this study, we aimed to develop an ECG-ASCVD model family, representing a comprehensive ECG-based risk prediction paradigm for predicting incident ASCVD within a diverse US health system. We then sought to validate its performance and compare it to PREVENT both internally and across two large volunteer cohorts in Brazil and the UK.

## Methods

### Data sources

Model development and internal validation were conducted using electronic health record (EHR) data from the Yale New Haven Health System (YNHHS), a multi-hospital health system serving a large, diverse population in the US. These data were linked to a state death registry to improve capture of cardiovascular mortality.^19^ Model performance was externally validated in two independent prospective cohort studies: the Brazilian Longitudinal Study of Adult Health (ELSA), a community-based prospective cohort of civil servants from Brazil, and the UK Biobank (UKB), a prospective observational volunteer cohort from the UK. Further information on data sources can be found in the **Supplemental Methods.** This study adheres to the TRIPOD+AI reporting guidelines.^20^ The Yale Institutional Review Board approved the study protocol and waived the need for informed consent, as the study involves analyzing preexisting data. Patients who opted out of research studies were excluded from the study analysis.

### Model development

We identified all individuals aged 30-79 in the YNHHS who had undergone a 12-lead ECG after 2013 and had at least 1 year of clinical data before and after the ECG. The earliest ECG on record was extracted for each individual, and its acquisition date was used as the study index date. Raw ECG voltage data were pre-processed using a standardized pipeline (Details in **Supplemental Methods**), which yielded 12-lead signals, 1-lead signals, and ECG images.

We trained 2 signal-based models, ECG-ASCVD-12 and ECG-ASCVD-1, which were one-dimensional residual convolutional neural networks (1D-ResNet) utilizing 12-lead signal information and information from only lead 1, respectively.^21^ Both models were first pre-trained as autoencoders to reconstruct ECG waveforms from Gaussian noise-augmented data to improve sample efficiency, robustness, and generalizability to wearable-based data.^22^

To develop our image-based model (ECG-ASCVD-IMAGE), we fine-tuned a BEiT-based vision transformer model, originally pretrained using contrastive learning on ECG images and corresponding echocardiogram reports.^23–25^ All pre-trained models were fitted with a multilayer perceptron head and fine-tuned to predict incident ASCVD using a neural network adapted time-to-event learning objective.^24^ We incorporated age and sex information into all models via decision fusion, where the final risk was computed as a linear combination of the ECG score and age and sex.

All individuals used in model pretraining were assigned to the fine-tuning training set. The remaining cohort was randomly split to yield training (70%), validation (10%), and test (20%) sets. Individuals with prevalent ASCVD were included in the training set as positive cases at time zero but were excluded from the test set. Hyperparameter optimization was conducted using the validation set, after which the best-performing model was retrained on all patients in the training and validation cohorts. Further details on input preprocessing, model architecture, and model training are specified in the **Supplemental Methods.**

### Study outcome and follow-up

The study outcome was time to first fatal or non-fatal occurrence of ASCVD. In the YNHHS, this was determined based on the presence of an inpatient clinical encounter or death record with a principal diagnosis of myocardial infarction (MI) or stroke using International Classification of Diseases, Tenth Revision Codes - Clinical Modification (ICD-10-CM) (specific codes are listed in **Supplementary Table S1**). The end of follow-up was defined as the date of death or the last recorded health contact in the EHR system. In ELSA, MI and stroke outcomes were identified through participant interviews with events verified by a centralized adjudication committee that systematically retrieved hospitalization records, outpatient reports, and death certificates.^26^ In the UKB self-reported outcomes and death certificate data were combined with ICD-10 codes extracted from a national EHR system.^27,28^

### Study covariates

PREVENT scores were calculated to serve as a comparison point for our models.^4^ We focused on the PREVENT-ASCVD models in their base form (without optional predictors of hemoglobin A1C, urine albumin creatinine ratio, or social deprivation index). In the YNHHS, predictor information was extracted from patient EHR records, encompassing all data from the 5 years preceding their index ECG. To account for delayed documentation, laboratory results and diagnoses recorded within 30 days after the index ECG were also included. Biochemical and biometric data were collected from inpatient and outpatient records. Where multiple results were available, the measure collected closest to the study index date was included.

In YNHHS, current smoking was defined based on a relevant ICD-10-CM diagnosis code, while diabetes mellitus was defined by either a diabetes-specific ICD-10-CM code or an HbA1c value greater than 6.5% (**Supplementary Table S2**). Blood pressure and lipid-lowering medication use were determined using outpatient prescription records and the Anatomical Therapeutic Chemical (ATC) classification system (**Supplementary Table S3**).^29^ In ELSA, diagnoses and medication use were self-reported, while biochemical and biometric variables were collected as part of the study protocol. In the UKB, current smoking and diabetes mellitus status were defined as in YNHHS, with the addition of patient self-reported information. Medications were identified from a nationally linked EHR and classified according to the ATC. Biochemical and biometric data were collected as part of the study protocol. Further information can be found in our previous paper and the **Supplemental Methods**.^30^

### Validation cohorts

Model performance was evaluated in individuals from the held-out testing set who also had (i) 1 year of information in the EHR system before their ECG, (ii) no recorded principal or secondary diagnosis of ASCVD before their index ECG. To compare our model against the PREVENT score, we also examined performance only in individuals with complete PREVENT variables, hereafter referred to as the YNHHS complete case cohort. In the UKB and ELSA, all individuals (i) aged 30-79, (ii) with an ECG, (iii) without prior ASCVD, and (iv) with complete PREVENT variables were included. Given protocolized testing, the external validation cohorts had higher rates of predictor availability, so only their complete case cohorts are used.

### Simulated deployment

To assess how these models might be deployed in a real-world clinical workflow, we examined the performance characteristics of ECG-ASCVD-1 and ECG-ASCVD-12 in individuals with at least 5 years of follow-up in the YNHHS held-out test set at an 80% sensitivity threshold. We then identified a sample of 100,000 randomly selected adults in YNHHS, examining the availability of PREVENT variables and clinical ECGs, and estimated the number of individuals who would screen positive at our selected threshold.

### Statistical analysis

Baseline characteristics were reported as counts and percentages for categorical variables and median and interquartile range (IQR) for continuous variables. ECG-ASCVD model discriminative performance for predicting incident ASCVD was assessed using Harrell’s C-index with 95% CIs computed using bootstrapping with 1,000 iterations.^31^

To assess whether ECG-ASCVD was an independent predictor for ASCVD over and above PREVENT, we used multivariate penalized Cox proportional hazards models that included age, sex, PREVENT, and ECG-ASCVD scores as covariates to calculate adjusted hazard ratios and associated p-values.

We used Python 3.11 and R version 4.2 to conduct all statistical analyses. All tests were two-sided with a significance level of 0.05. Our analysis code and fine-tuned model weights are published online for research use at https://github.com/CarDS-Yale/ecg-ascvd and https://huggingface.co/CarDSLab/.

## Results

### Study populations

We identified 454,736 unique individuals with an eligible ECG at YNHHS (**Supplemental Figure 1**). The development population (comprising model training and validation) consisted of 363,788 individuals with a median age of 57.1 years (Q1-Q3: 45.5-67.2 years). In this population, 174,940 (48.1%) were women, 254,017 (69.8%) were white, and 17,345 (4.8%) experienced an ASCVD event over a median 6.9 years of follow-up (**Table 1**).

**Table 1:**
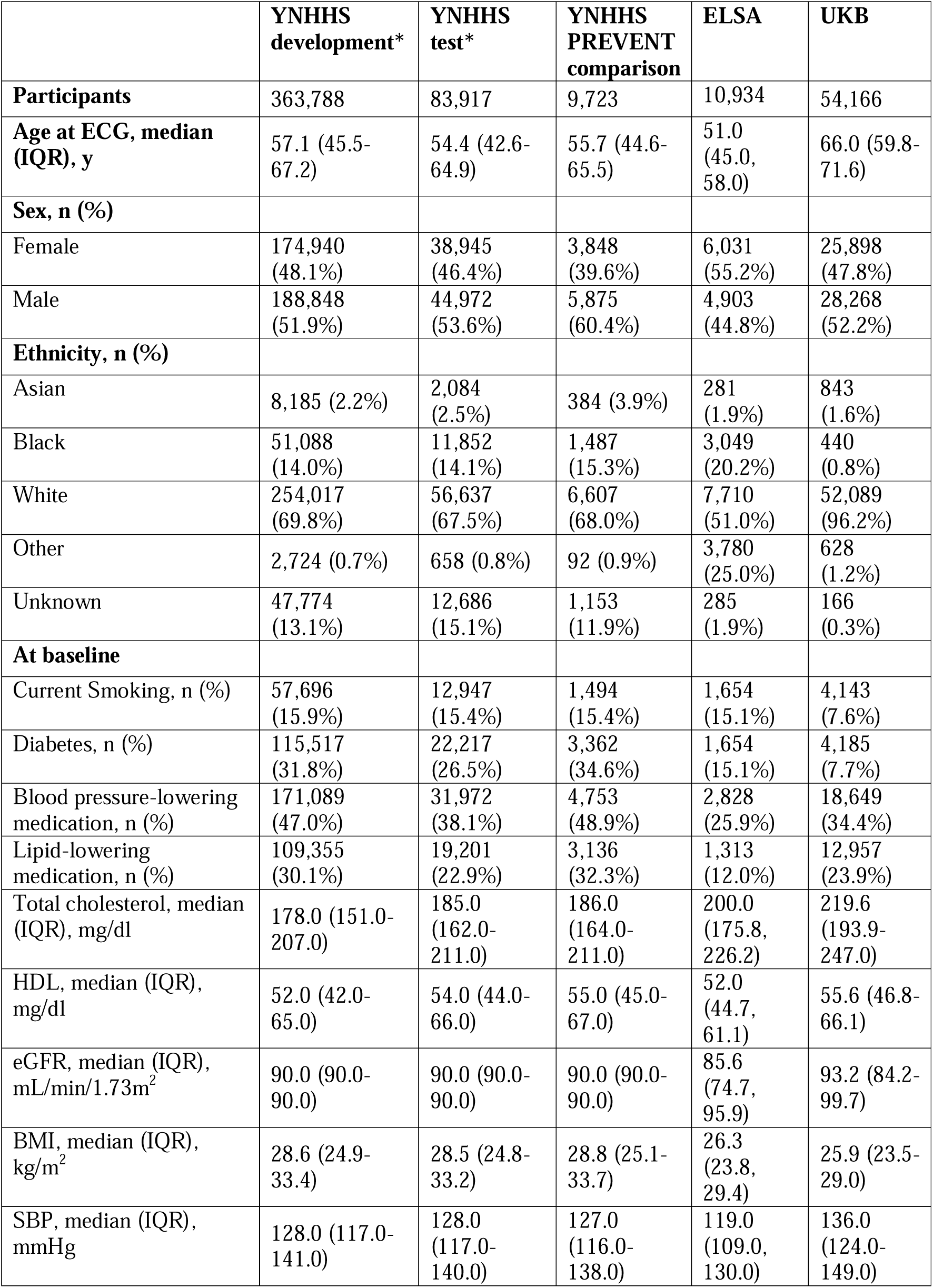
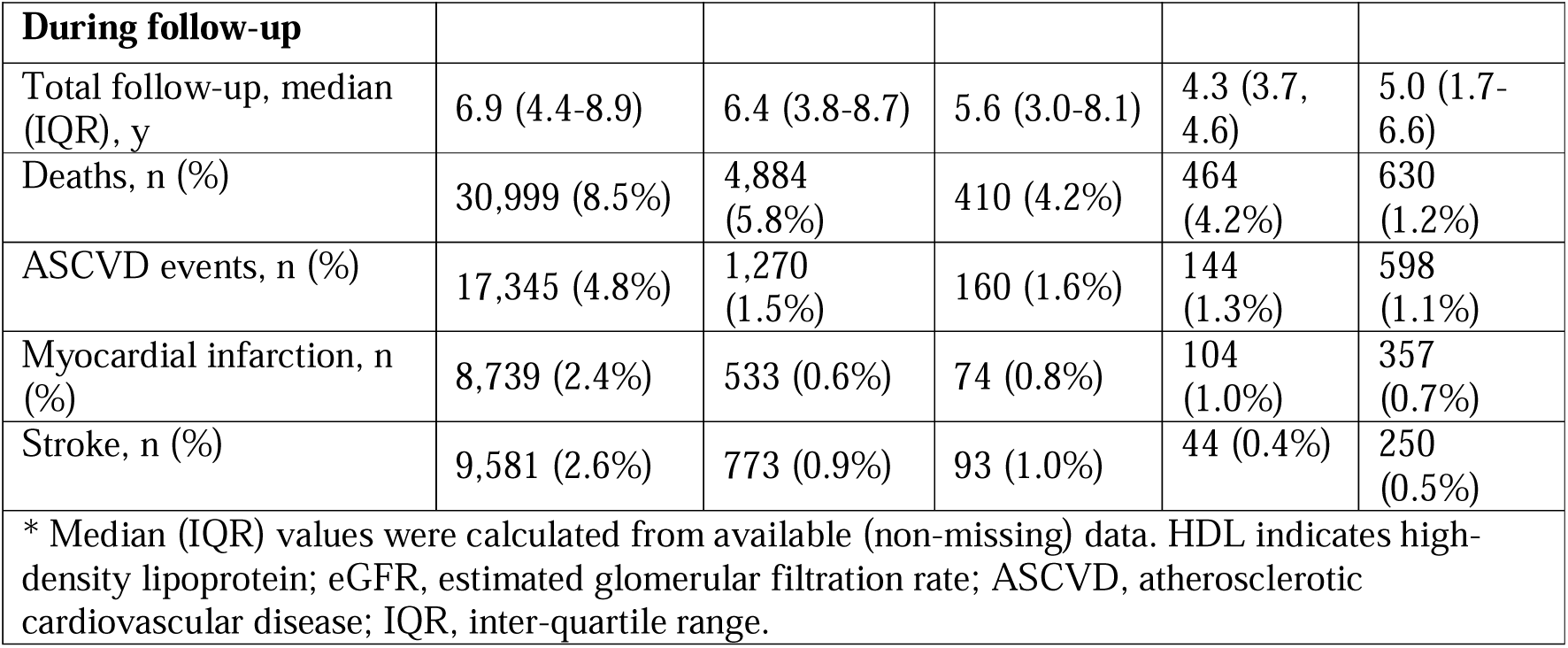
Demographic characteristics of the study cohorts.

|  | **YNHHS**<br>**development\*** | **YNHHS**<br>**test\*** | **YNHHS**<br>**PREVENT**<br>**comparison** | **ELSA** | **UKB** |
| --- | --- | --- | --- | --- | --- |
| **Participants** | 363,788 | 83,917 | 9,723 | 10,934 | 54,166 |
| **Age at ECG, median (IQR), y** | 57.1 (45.5-67.2) | 54.4 (42.6-64.9) | 55.7 (44.6-65.5) | 51.0 (45.0, 58.0) | 66.0 (59.8-71.6) |
| **Sex, n (%)** |  |  |  |  |  |
| Female | 174,940 (48.1%) | 38,945 (46.4%) | 3,848 (39.6%) | 6,031 (55.2%) | 25,898 (47.8%) |
| Male | 188,848 (51.9%) | 44,972 (53.6%) | 5,875 (60.4%) | 4,903 (44.8%) | 28,268 (52.2%) |
| **Ethnicity, n (%)** |  |  |  |  |  |
| Asian | 8,185 (2.2%) | 2,084 (2.5%) | 384 (3.9%) | 281 (1.9%) | 843 (1.6%) |
| Black | 51,088 (14.0%) | 11,852 (14.1%) | 1,487 (15.3%) | 3,049 (20.2%) | 440 (0.8%) |
| White | 254,017 (69.8%) | 56,637 (67.5%) | 6,607 (68.0%) | 7,710 (51.0%) | 52,089 (96.2%) |
| Other | 2,724 (0.7%) | 658 (0.8%) | 92 (0.9%) | 3,780 (25.0%) | 628 (1.2%) |
| Unknown | 47,774 (13.1%) | 12,686 (15.1%) | 1,153 (11.9%) | 285 (1.9%) | 166 (0.3%) |
| **At baseline** |  |  |  |  |  |
| Current Smoking, n (%) | 57,696 (15.9%) | 12,947 (15.4%) | 1,494 (15.4%) | 1,654 (15.1%) | 4,143 (7.6%) |
| Diabetes, n (%) | 115,517 (31.8%) | 22,217 (26.5%) | 3,362 (34.6%) | 1,654 (15.1%) | 4,185 (7.7%) |
| Blood pressure-lowering medication, n (%) | 171,089 (47.0%) | 31,972 (38.1%) | 4,753 (48.9%) | 2,828 (25.9%) | 18,649 (34.4%) |
| Lipid-lowering medication, n (%) | 109,355 (30.1%) | 19,201 (22.9%) | 3,136 (32.3%) | 1,313 (12.0%) | 12,957 (23.9%) |
| Total cholesterol, median (IQR), mg/dl | 178.0 (151.0-207.0) | 185.0 (162.0-211.0) | 186.0 (164.0-211.0) | 200.0 (175.8, 226.2) | 219.6 (193.9-247.0) |
| HDL, median (IQR), mg/dl | 52.0 (42.0-65.0) | 54.0 (44.0-66.0) | 55.0 (45.0-67.0) | 52.0 (44.7, 61.1) | 55.6 (46.8-66.1) |
| eGFR, median (IQR), mL/min/1.73m<sup>2</sup> | 90.0 (90.0-90.0) | 90.0 (90.0-90.0) | 90.0 (90.0-90.0) | 85.6 (74.7, 95.9) | 93.2 (84.2-99.7) |
| BMI, median (IQR), kg/m<sup>2</sup> | 28.6 (24.9-33.4) | 28.5 (24.8-33.2) | 28.8 (25.1-33.7) | 26.3 (23.8, 29.4) | 25.9 (23.5-29.0) |
| SBP, median (IQR), mmHg | 128.0 (117.0-141.0) | 128.0 (117.0-140.0) | 127.0 (116.0-138.0) | 119.0 (109.0, 130.0) | 136.0 (124.0-149.0) |
| **During follow-up** |  |  |  |  |  |
| Total follow-up, median (IQR), y | 6.9 (4.4-8.9) | 6.4 (3.8-8.7) | 5.6 (3.0-8.1) | 4.3 (3.7, 4.6) | 5.0 (1.7-6.6) |
| Deaths, n (%) | 30,999 (8.5%) | 4,884 (5.8%) | 410 (4.2%) | 464 (4.2%) | 630 (1.2%) |
| ASCVD events, n (%) | 17,345 (4.8%) | 1,270 (1.5%) | 160 (1.6%) | 144 (1.3%) | 598 (1.1%) |
| Myocardial infarction, n (%) | 8,739 (2.4%) | 533 (0.6%) | 74 (0.8%) | 104 (1.0%) | 357 (0.7%) |
| Stroke, n (%) | 9,581 (2.6%) | 773 (0.9%) | 93 (1.0%) | 44 (0.4%) | 250 (0.5%) |
| * Median (IQR) values were calculated from available (non-missing) data. HDL indicates high-density lipoprotein; eGFR, estimated glomerular filtration rate; ASCVD, atherosclerotic cardiovascular disease; IQR, inter-quartile range. |  |  |  |  |  |

The held-out test set included 83,917 individuals (median age 54.4, Q1-Q3: 42.6-64.9) without prior ASCVD, of whom 38,945 (46.4%) were women, 56,637 (67.5%) were white, and 1,270 (1.5%) experienced an ASCVD event during 6.4 years of follow-up (Q1-Q3: 3.8-8.7). Only 9,723 (11.6%) of the testing set had all PREVENT variable information extractable from their EHR prior to their ECG.

The external validation cohorts encompassed 10,934 and 54,166 individuals without prior ASCVD and with complete PREVENT variable information in ELSA and the UKB, respectively (**Supplementary Figure S2-3**). Individuals in ELSA were younger (median 51.0, Q1-Q3: 45.0-58.0), while those in the UKB were older (median 66.0, Q1-Q3: 59.8-71.6) than the model development population. ASCVD rates were similar in both cohorts, with 144 (1.3%) and 598 (1.1%) individuals experiencing ASCVD over 4.3 years (IQR: 3.7-4.6) and 5.0 years (IQR: 1.7-6.6) of follow-up in ELSA and the UKB, respectively (**Figure 1**).

**Figure 1:**
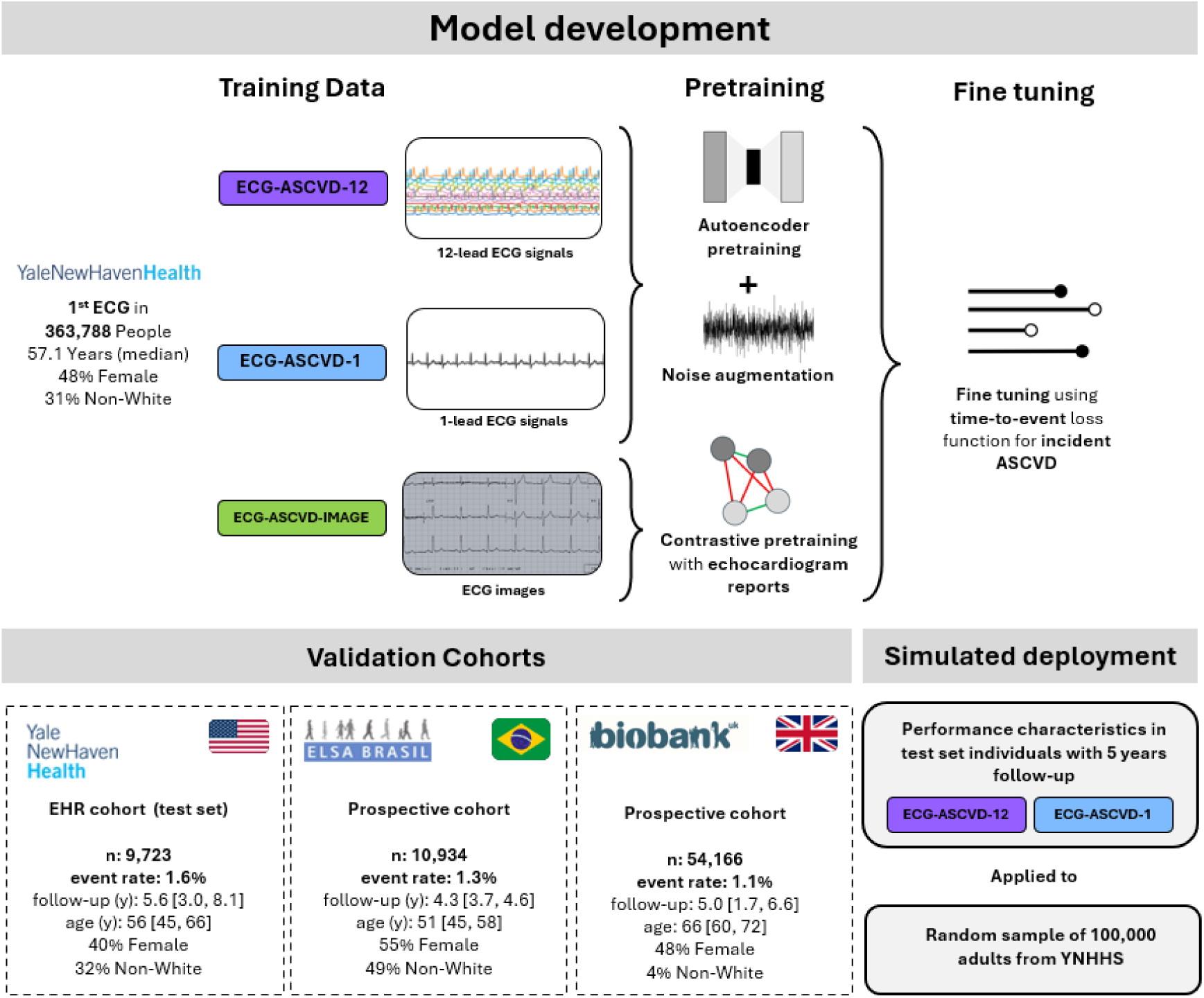
Study Overview.

### Model performance

Across the YNHHS test set, all three ECG-ASCVD models demonstrated largely equivalent discriminative performance, with concordance values of 0.709 (95% CI, 0.698-0.721) for ECG-ASCVD-1, 0.711 (95% CI, 0.699-0.722) for ECG-ASCVD-12, and 0.714 (95% CI, 0.703-0.726) for ECG-ASCVD-IMAGE (**Figure 2**). In the YNHHS complete case cohort, ECG-ASCVD-12 achieved a C-index of 0.723 (0.691-0.756), closely tracking the PREVENT C-index of 0.724 (0.689-0.758). Performance was also comparable in the UKB, with C-indexes of 0.684 (0.665-0.704) and 0.696 (0.678-0.714), respectively. However, ECG-ASCVD-12 was inferior in ELSA-Brasil, reaching a C-index of 0.746 (0.706-0.786) compared with 0.782 (0.746-0.817) for PREVENT (**Supplementary Table 4**).

**Figure 2:**
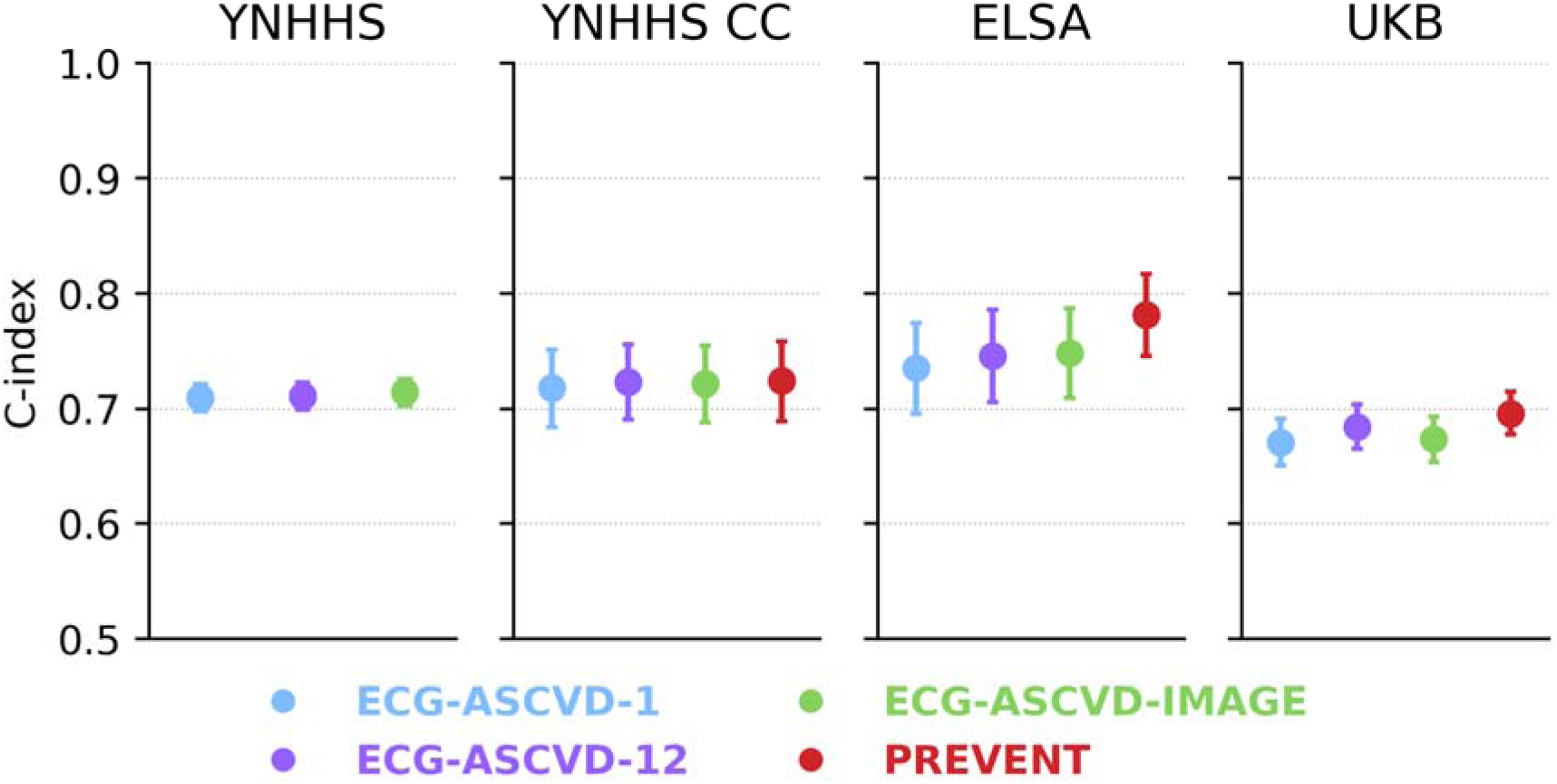
ECG-ASCVD model performance in the YNHHS test set, YNHHS prevent comparison set, ELSA, and UKB. YNHHS CC indicates Yale New Haven Hospital System complete cases; ELSA, Brazilian Longitudinal Study of Adult Health; UKB, UK Biobank; PREVENT, Predicting Risk of Cardiovascular Disease EVENTs equations.

ECG-ASCVD-IMAGE showed performance comparable to ECG-ASCVD-12, with C-indices of 0.721 (95% CI, 0.688-0.755), 0.748 (0.709-0.787), and 0.673 (0.653-0.694) in the YNHHS complete case cohort, ELSA, and the UKB, respectively. ECG-ASCVD-1 demonstrated modestly lower discrimination, with corresponding C-indices of 0.718 (0.684-0.751), 0.735 (0.696-0.774), and 0.671 (0.651-0.691) in the same cohorts, respectively. In the prediction of 10-year ASCVD incidence, all model calibration closely mirrored PREVENT. The models were well calibrated in the YNHHS, but overestimated risk in the ELSA and UKB cohorts (**Supplementary Figure S4**).

### Additive benefit over PREVENT

Across all three cohorts, the ECG-ASCVD-12 score demonstrated an independent association with incident ASCVD beyond the PREVENT score. In penalized Cox models (ridge penalty, λ = 0.01) including age, sex, ECG-ASCVD-12, and PREVENT, the ECG-based score remained a significant predictor with adjusted hazard ratios of 1.34 (95% CI 1.12-1.51) in the YNHHS complete-case sample, 1.23 (95% CI 1.04-1.59) in ELSA, and 1.33 (95% CI 1.20-1.48) in UKB (**Figure 3** and **Supplementary Table 5).**

**Figure 3:**
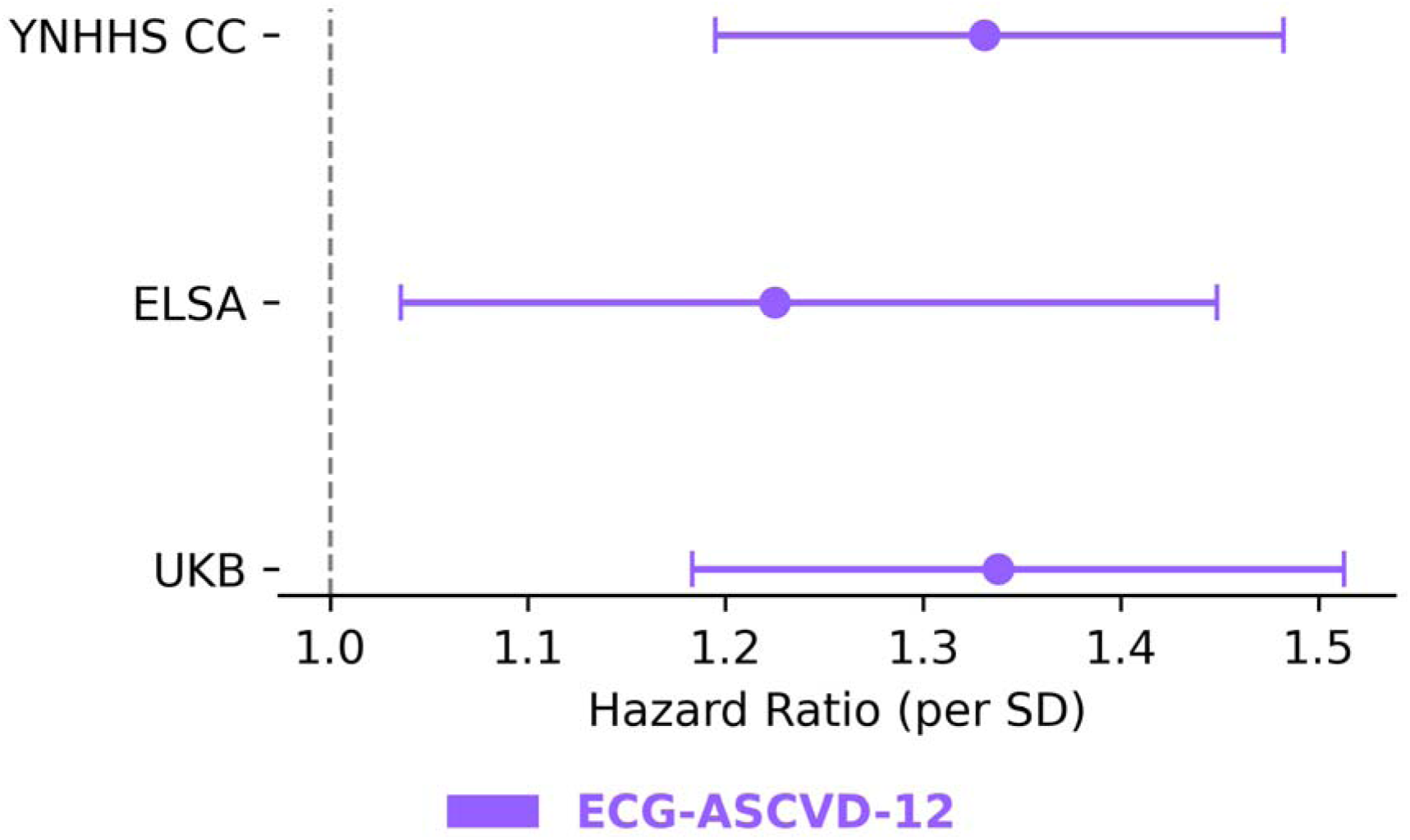
Adjusted hazard ratios of the ECG-ASCVD-12 score, controlling for PREVENT, age, and sex in the 3 PREVENT comparison cohorts, for predicting ASCVD. YNHHS CC indicates Yale New Haven Hospital System complete cases; ELSA, Brazilian Longitudinal Study of Adult Health; UKB, UK Biobank.

### Simulated deployment

Of 100,000 randomly selected adults identified in the YNHHS, 72,654 were between the ages of 30 and 79, and 70,476 of these individuals had no prior history of ASCVD and were thus eligible to be risk-assessed by PREVENT. Of this PREVENT eligible population, only 12.5% had all the necessary predictor information in their EHR to calculate a PREVENT score, while a further 16.5% had a clinical ECG but insufficient information to calculate the PREVENT score (**Figure 4**). Total cholesterol and HDL cholesterol measures were the most frequently missing predictors, with only 18.9% of individuals having a test recorded. The missingness of other predictors is detailed in **Supplementary Table 6.**

**Figure 4:**
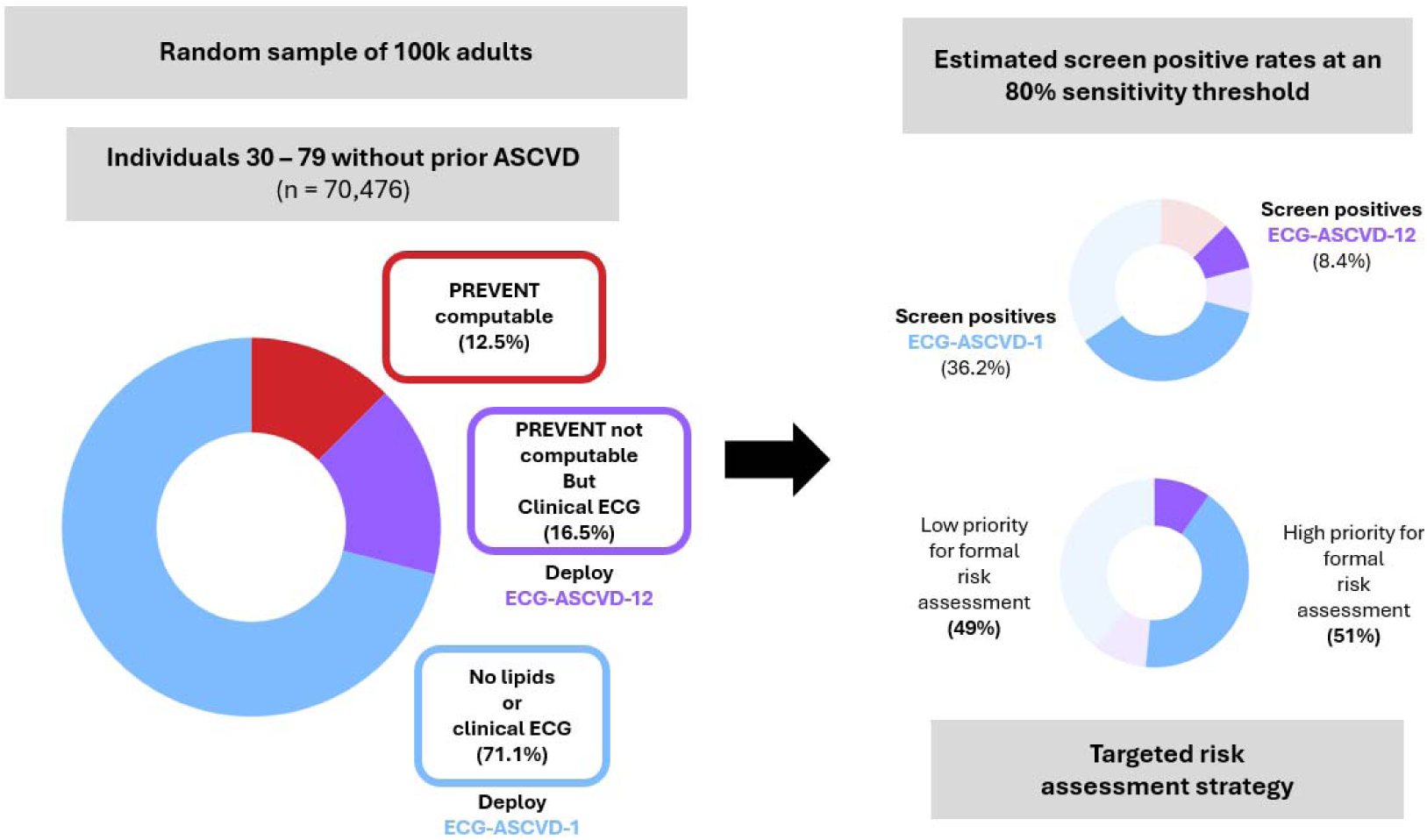
Simulated deployment of ECG-ASCVD-12 and ECG-ASCVD-1 in a random sample of 100k adults to help target risk factor assessment. PREVENT indicates Predicting Risk of Cardiovascular Disease EVENTs equations.

In a subset of the YNHHS test set with at least 5 years of follow-up, ECG-ASCVD-1 and ECG-ASCVD-12 both had a screen-positive proportion of 0.51 at an 80% sensitivity threshold. Therefore, if the ECG-ASCVD-12 model was run on all clinical ECGs among individuals without a computable PREVENT score, and the ECG-ASCVD-1 model was run on all individuals without either PREVENT or a clinical ECG, we estimate that 51% of the population in whom 80% of events would occur over 5 years would flag positive.

## Discussion

In this large, multinational study, we developed and validated ECG-ASCVD, a deep learning-based ASCVD risk prediction paradigm that utilizes 12-lead ECG signals, ECG-images, and 1-lead ECG signals to aid in targeting risk factor assessment. ECG-ASCVD-12 demonstrated generalizable discriminative performance in predicting the first onset of ASCVD with C-indices ranging from 0.684 to 0.746 in a held-out real-world US health-system test cohort and across two international prospective cohorts representing diverse middle-income (ELSA) and high-income (UKB) populations. An image-based model, ECG-ASCVD-IMAGE, was almost equivalent (C-indices ranging from 0.673 to 0.748) to one trained on ECG signal data, with both approaching the discriminative performance of PREVENT (C-indices ranging from 0.696 to 0.782). A model that utilized lead-1 signal data exhibited only modestly attenuated performance (C-indices ranging from 0.671 to 0.735), while having the potential to be applied to increasingly common ECG-enabled wearable and portable devices. ECG-ASCVD could identify individuals who account for the majority of ASCVD events yet lack consistent risk factor assessment needed to compute traditional risk scores.

Prior studies have demonstrated that structured ECG parameters carry prognostic signals for ASCVD, and more recently, deep learning approaches have been applied to raw waveform data, improving model performance.^11,16–18^ However, these approaches rely on access to raw waveform data and specialized preprocessing, which limits their scalability and uptake, especially in resource-constrained settings where they would be the most useful.^16,18^. Our study is the first to develop and validate an ECG image-based deep learning model for ASCVD prediction, demonstrating robust performance across diverse populations and achieving comparable performance to waveform-based models.^18^ Our study further supports previous work by our team and others, suggesting that for several cardiac diseases, waveform and image-based ECG models are generally equivalent in predictive performance.^12,32^

For the prediction of ASCVD, the performance of the AI-ECG models has so far only been compared against the Pooled Cohort Equations (PCEs), which have now been superseded in practice by the PREVENT equations.^33^ The PREVENT equations, introduced by the American Heart Association, were created to address issues arising from the inclusion of race as a feature in PCEs and to add more complete metabolic risk factor information. Our study is the first to directly compare an AI-ECG score with the PREVENT equations and demonstrate that the AI-ECG score is an independent risk factor for ASCVD, even when using a risk score that incorporates cardiometabolic information.

Our study was also the first to examine the utility of lead-1 ECG data, which can now be captured by ECG-enabled wearable and portable devices, to predict ASCVD. While a modest drop in discriminative performance was observed in the external validation cohorts, ECG-ASCVD-1 holds the potential for widespread deployment outside traditional clinical settings. Given the tendency of wearable devices to capture noisier signals, we employed an artificial augmentation of the lead-1 ECGs during model pre-training to improve robustness to signals captured from wearable/portable devices.^22^ Targeted validation using ECGs captured from these devices is a crucial next step before deploying such models clinically.

Our findings suggest three possible applications for ECG-ASCVD. First, parsimonious risk assessments could be conducted in low-resource settings where laboratory testing is not broadly available or feasible for ASCVD risk assessment. The rise of open-source ECG platforms and consumer wearable device technology has rapidly driven down the cost of acquiring ECG data, allowing image or single-lead models to be easily deployed via a web app or smartphone, thereby reaching more resource-deprived areas. ^34,35^ Second, given the lack of risk assessments being conducted even among the selected population of individuals who have received an ECG at YNHHS (only 18% of adults having a total cholesterol test in their EHR record, and only 13% having all the variables required for PREVENT), the ECG-ASCVD score could be used to help pre-screen individuals, identifying who would benefit most from a formal risk assessment. Our models could be applied passively in the EHR and nudge clinicians caring for patients with high ECG-ASCVD scores to consider formal ASCVD risk assessment, or they can be widely deployed in community settings through wearable devices.^36^

Lastly, the adjusted hazard ratios of the ECG-ASCVD score, when controlling for PREVENT, indicate the ability of ECG-ASCVD to alter individual risk predictions.^37^ The PREVENT equations offer several optional variables (HbA1c, UACR, ZIP code) that individuals can include to refine their risk assessment.^4^ An AI-ECG risk score could thus be another variable offered, with clinicians uploading a picture of a patient’s ECG, to refine individual-level risk assessment.

Our work, along with the growing number of studies demonstrating the prognostic value of resting ECG information, challenges current guidelines that do not recommend resting ECGs in asymptomatic individuals.^38^ While prospective and randomized clinical trial data are required before ECG models are used to inform care, routine ECG screening could become similar to blood pressure measurement (primarily of interest due to its role as a risk factor for ASCVD), which is now ubiquitous.

This study has limitations that merit consideration. First, we identified ASCVD events in YNHHS solely through ICD-10 codes, which were recorded in the EHR, and ICD-10-coded causes of death in a state death registry. This fails to capture information stored in procedure codes (e.g., revascularization) and biochemical tests (e.g., troponin elevation), introducing systematic variability in the outcome measure used to train our model.^39^ However, this design choice purposely mirrored the methodology used to derive and validate the PREVENT equations in electronic health record (EHR) settings.^4^ Furthermore, linkage to the state death registry broadened capture of fatal events and partly mitigated missed out-of-hospital presentations. Second, risk-factor measurements were obtained retrospectively from routine care and therefore did not exactly coincide with an individual’s study index date. Temporal dispersion increases within-person biological variability, potentially diluting regression coefficients and modestly lowering discrimination. Nevertheless, the PREVENT C-index observed in our complete-case YNHHS cohort closely matches the performance reported in other studies that have examined similar cohorts.^4,30,40–42^ Third, the drop in performance observed in the UKB cohort compared to ELSA and YNHHS may signal model overfitting. However, a similar drop in discriminative performance has been noted in other studies that have externally validated ECG prognostic models in the UKB.^18^ The notable differences in the C-index of the PREVENT equations model further suggest that this change in model C-index likely reflects genuine between-cohort differences in case mix and risk structure.^35^

## Conclusion

We developed and validated a novel ECG-based ASCVD risk assessment toolkit, deployable on both clinical 12-lead ECG images and portable/wearable 1-lead ECGs, across clinically and geographically diverse datasets. ECG-ASCVD represents a pragmatic, scalable, and accessible strategy for identifying patients who would most benefit from formal risk assessment, thereby supporting the clinical potential of routine ECG-based screening.

## Data Sharing Statement

Data from the UK Biobank and the Brazilian Longitudinal Study of Adult Health are available for research to licensed users. Individual-level data for the Yale New Haven Health System cannot be made available due to HIPAA regulations enforced by the Yale IRB. The models presented here are available for research use by request on our website and on Hugging Face (https://huggingface.co/CarDSLab/ECG-ASCVD-1, https://huggingface.co/CarDSLab/ECG-ASCVD-12, and https://huggingface.co/CarDSLab/ECG-ASCVD-IMAGE), and code for generating key results is available in a public GitHub repository (https://github.com/CarDS-Yale/ecg-ascvd).

## Author contributions

B.B and R.K designed the study. B.B, A.P, L.D, and A.A performed data cleaning and pre-processing. B.B and E.O implemented the models. B.B evaluated model performance and drafted the manuscript. All authors contributed to the interpretation of findings and manuscript revisions.

## References

1. Daalen KR van, Zhang D, Kaptoge S, Paige E, Angelantonio ED, Pennells L. Risk estimation for the primary prevention of cardiovascular disease: considerations for appropriate risk prediction model selection. The Lancet Global Health. 2024;12(8):e1343–e1358. doi:10.1016/S2214-109X(24)00210-9

2. Damen JAAG, Hooft L, Schuit E, et al. Prediction models for cardiovascular disease risk in the general population: systematic review. BMJ. Published online 2016:i2416. doi:10.1136/bmj.i2416

3. Shillinglaw B, Viera AJ, Edwards T, Simpson R, Sheridan SL. Use of global coronary heart disease risk assessment in practice: a cross-sectional survey of a sample of U.S. physicians. BMC Health Serv Res. 2012;12(1):20. doi:10.1186/1472-6963-12-20

4. Khan SS, Matsushita K, Sang Y, et al. Development and Validation of the American Heart Association’s PREVENT Equations. Circulation. 2024;149(6):430–449. doi:10.1161/circulationaha.123.067626

5. Finnikin S, Willis BH, Ryan R, Evans T, Marshall T. Factors predicting statin prescribing for primary prevention: a historical cohort study. Br J Gen Pract. 2021;71(704):e219–e225. doi:10.3399/bjgp20X714065

6. Miedema MD, Garberich RF, Schnaidt LJ, et al. Statin Eligibility and Outpatient Care Prior to STDSegment Elevation Myocardial Infarction. J Am Heart Assoc. 2017;6(4):e005333. doi:10.1161/JAHA.116.005333

7. Mensah GA, Fuster V, Murray CJL, et al. Global Burden of Cardiovascular Diseases and Risks, 1990-2022. Journal of the American College of Cardiology. 2023;82(25):2350–2473. doi:10.1016/j.jacc.2023.11.007

8. Reichlin T, Abächerli R, Twerenbold R, et al. Advanced ECG in 2016: is there more than just a tracing? Swiss Medical Weekly. 2016;146(1718):w14303–w14303. doi:10.4414/smw.2016.14303

9. Rjoob K, Bond R, Finlay D, et al. Machine learning and the electrocardiogram over two decades: Time series and meta-analysis of the algorithms, evaluation metrics and applications. Artif Intell Med. 2022;132:102381. doi:10.1016/j.artmed.2022.102381

10. Zvuloni E, Read J, Ribeiro AH, Ribeiro ALP, Behar JA. On Merging Feature Engineering and Deep Learning for Diagnosis, Risk Prediction and Age Estimation Based on the 12-Lead ECG. IEEE Trans Biomed Eng. 2023;70(7):2227–2236. doi:10.1109/TBME.2023.3239527

11. Shah AJ, Vaccarino V, Janssens ACJW, et al. An Electrocardiogram-Based Risk Equation for Incident Cardiovascular Disease From the National Health and Nutrition Examination Survey. JAMA Cardiology. 2016;1(7):779–786. doi:10.1001/jamacardio.2016.2173

12. Dhingra LS, Aminorroaya A, Sangha V, et al. Ensemble Deep Learning Algorithm for Structural Heart Disease Screening Using Electrocardiographic Images: PRESENT SHD. Journal of the American College of Cardiology. 2025;85(12):1302–1313. doi:10.1016/j.jacc.2025.01.030

13. Raghunath S, Ulloa Cerna AE, Jing L, et al. Prediction of mortality from 12-lead electrocardiogram voltage data using a deep neural network. Nat Med. 2020;26(6):886–891. doi:10.1038/s41591-020-0870-z

14. Lima EM, Ribeiro AH, Paixão GMM, et al. Deep neural network-estimated electrocardiographic age as a mortality predictor. Nat Commun. 2021;12(1):5117. doi:10.1038/s41467-021-25351-7

15. Sau A, Barker J, Pastika L, et al. Artificial Intelligence–Enhanced Electrocardiography for Prediction of Incident Hypertension. JAMA Cardiol. 2025;10(3):214. doi:10.1001/jamacardio.2024.4796

16. Hughes JW, Tooley J, Torres Soto J, et al. A deep learning-based electrocardiogram risk score for long term cardiovascular death and disease. npj Digit Med. 2023;6(1):1–9. doi:10.1038/s41746-023-00916-6

17. Awasthi S, Sachdeva N, Gupta Y, et al. Identification and risk stratification of coronary disease by artificial intelligence-enabled ECG. eClinicalMedicine. 2023;65. doi:10.1016/j.eclinm.2023.102259

18. Sau A, Pastika L, Sieliwonczyk E, et al. Artificial intelligence-enabled electrocardiogram for mortality and cardiovascular risk estimation: a model development and validation study. The Lancet Digital Health. 2024;6(11):e791–e802. doi:10.1016/S2589-7500(24)00172-9

19. CT-Vitals Help. Accessed July 22, 2025. https://ctvitals.ct.gov/

20. TRIPOD+AI statement: updated guidance for reporting clinical prediction models that use regression or machine learning methods | The BMJ. Accessed August 12, 2025. https://www.bmj.com/content/385/bmj-2023-078378

21. He K, Zhang X, Ren S, Sun J. Deep Residual Learning for Image Recognition. In: 2016 IEEE Conference on Computer Vision and Pattern Recognition (CVPR). 2016:770–778. doi:10.1109/CVPR.2016.90

22. Khunte A, Sangha V, Oikonomou EK, et al. Detection of left ventricular systolic dysfunction from single-lead electrocardiography adapted for portable and wearable devices. NPJ Digit Med. 2023;6(1):124. doi:10.1038/s41746-023-00869-w

23. Bao H, Dong L, Piao S, Wei F. BEiT: BERT Pre-Training of Image Transformers. arXiv. Preprint posted online September 3, 2022. doi:10.48550/arXiv.2106.08254

24. Monod M, Krusche P, Cao Q, et al. TorchSurv: A Lightweight Package for Deep Survival Analysis. JOSS. 2024;9(104):7341. doi:10.21105/joss.07341

25. Oikonomou EK, Batinica B, Dhingra LS, Aminorroaya A, Coppi A, Khera R. TARGET-AI: a foundational approach for the targeted deployment of artificial intelligence electrocardiography in the electronic health record. medRxiv. Published online October 25, 2025:2025.08.25.25334266. doi:10.1101/2025.08.25.25334266

26. Barreto SM, Ladeira RM, Bastos MDSCBDO, et al. Estratégias de identificação, investigação e classificação de desfechos incidentes no ELSA-Brasil. Rev Saúde Pública. 2013;47(suppl 2):79–86. doi:10.1590/s0034-8910.2013047003836

27. Rubbo B, Fitzpatrick NK, Denaxas S, et al. Use of electronic health records to ascertain, validate and phenotype acute myocardial infarction: A systematic review and recommendations. International Journal of Cardiology. 2015;187:705–711. doi:10.1016/j.ijcard.2015.03.075

28. Schnier C, Denaxas S, Eggo R, et al. Identification and validation of myocardial infarction and stroke outcomes at scale in UK Biobank: IJPDS (2017) Issue 1, Vol 1:337 Proceedings of the IPDLN Conference (August 2016). IJPDS. 2017;1(1). doi:10.23889/ijpds.v1i1.358

29. ATCDDD - ATC/DDD Index. Accessed July 21, 2025. https://atcddd.fhi.no/atc_ddd_index/

30. Camargos AP, Barreto S, Brant L, et al. Performance of contemporary cardiovascular risk stratification scores in Brazil: an evaluation in the ELSA-Brasil study. Open Heart. 2024;11(1). doi:10.1136/openhrt-2024-002762

31. Harrell, FE. Regression Modeling Strategies: With Applications to Linear Models, Logistic and Ordinal Regression, and Survival Analysis. Springer International Publishing; 2015. doi:10.1007/978-3-319-19425-7

32. Sau A, Zeidaabadi B, Patlatzoglou K, et al. A comparison of artificial intelligence–enhanced electrocardiography approaches for the prediction of time to mortality using electrocardiogram images. European Heart Journal - Digital Health. 2025;6(2):180–189. doi:10.1093/ehjdh/ztae090

33. Goff DC, Lloyd-Jones DM, Bennett G, et al. 2013 ACC/AHA Guideline on the Assessment of Cardiovascular Risk: A Report of the American College of Cardiology/American Heart Association Task Force on Practice Guidelines. Circulation. 2014;129(25_suppl_2). doi:10.1161/01.cir.0000437741.48606.98

34. Ertola JP, Figueira S, Carlsen M, Palaniappan U, Rondini K. Low-cost electrocardiogram device for preventive health care in rural populations of developing countries. In: 2016 IEEE Global Humanitarian Technology Conference (GHTC). 2016:646–655. doi:10.1109/GHTC.2016.7857347

35. Wang R, Veera SCM, Asan O, Liao T. A Systematic Review on the Use of Consumer-Based ECG Wearables on Cardiac Health Monitoring. IEEE Journal of Biomedical and Health Informatics. 2024;28(11):6525–6537. doi:10.1109/JBHI.2024.3456028

36. Ghazi L, Yamamoto Y, Riello RJ, et al. Electronic Alerts to Improve Heart Failure Therapy in Outpatient Practice. JACC. 2022;79(22):2203–2213. doi:10.1016/j.jacc.2022.03.338

37. Cook NR. Use and Misuse of the Receiver Operating Characteristic Curve in Risk Prediction. Circulation. 2007;115(7):928–935. doi:10.1161/CIRCULATIONAHA.106.672402

38. US Preventive Services Task Force. Screening for Cardiovascular Disease Risk With Electrocardiography: US Preventive Services Task Force Recommendation Statement. JAMA. 2018;319(22):2308–2314. doi:10.1001/jama.2018.6848

39. Bosco E, Hsueh L, McConeghy KW, Gravenstein S, Saade E. Major adverse cardiovascular event definitions used in observational analysis of administrative databases: a systematic review. BMC Med Res Methodol. 2021;21(1):241. doi:10.1186/s12874-021-01440-5

40. Singh A, Shiyovich A, Freire CV, et al. Performance of PREVENT equations for cardiovascular risk prediction in young patients with myocardial infarction: From the MGB YOUNG-MI registry. American Journal of Preventive Cardiology. 2025;22:100992. doi:10.1016/j.ajpc.2025.100992

41. Cho SMJ, Levin M, Chen R, et al. AHA PREVENT Equations and Cardiovascular Disease Risk in Diverse Health Care Populations. Journal of the American College of Cardiology. 2025;86(3):181–192. doi:10.1016/j.jacc.2025.04.066

42. Ambrosio M, Alebna PL, Lee T, et al. Performance of PREVENT and pooled cohort equations for predicting 10-Year ASCVD risk in the UK Biobank. American Journal of Preventive Cardiology. 2025;22:101009. doi:10.1016/j.ajpc.2025.101009

