## Supplement for "Development and Multinational Validation of Artificial Intelligence-Enabled ASCVD Risk Stratification Using Electrocardiograms"

### SUPPLEMENTARY MATERIALS

### SUPPLEMENTAL METHODS:

#### Data sources and study population:

Electronic health record (EHR) data were obtained for patients receiving care within the Yale New Haven Health System between 2013 and 2024. YNHHS comprises eight hospitals and serves as the sole provider for a demographically diverse patient population representing approximately one-third of the state of Connecticut. Clinical data were recorded in the Epic EHR platform and extracted from the Epic Clarity database. Mortality data occurring outside the hospital system were incorporated through linkage with the Connecticut death index (CTVitals). For the present study, electrocardiograms (ECGs) performed within one month of a cardiovascular disease (CVD) event were excluded from analysis.

The UK Biobank is a population-based prospective cohort that recruited 502,468 community-dwelling adults aged 40-69 years between 2006 and 2010.^1^ A subset of participants attended a third or fourth follow-up visit between 2014 and 2021, during which 12-lead ECGs were obtained. UKB data are linked to the United Kingdom’s National Health Service EHR, providing diagnosis and procedure codes, including those predating study enrollment, and to the national death registry for comprehensive ascertainment of mortality.^2^ Data were accessed under research application #71033.

ELSA-Brasil is a multicenter, prospective study investigating the determinants and progression of chronic diseases among Brazilian adults. Between 2008 and 2010, 15,105 active and retired civil servants aged 35-74 years were enrolled from six public universities and research institutions in three regions: Southeast (Belo Horizonte, Rio de Janeiro, São Paulo, Vitória), South (Porto Alegre), and Northeast (Salvador).^3,4^ Baseline evaluations included validated questionnaires, physical examinations, laboratory analyses, and imaging modalities such as 12-lead ECG and echocardiography.^4^ Follow-up assessments occur every 3-4 years in person, supplemented by annual telephone interviews to document incident diagnoses, hospitalizations, and deaths.^4^ Reported clinical events undergo adjudication by expert review of medical records.

#### PREVENT variable extraction in YNHHS, UKB, and ELSA:

In the YNHHS, predictor information was extracted retrospectively from individual EHR records. Current smoking was defined based on the presence of an ICD-10 code for smoking (Z72, Z87, F17) in the last year prior to study inclusion. Diabetes was defined based on ICD-10 codes (E11) or an HbA1c results of >6.5%. Systolic blood pressure results were extracted from the most recent set of vitals taken prior to study inclusion. Extreme lab values (outside the range of the prevent calculator) were clamped to the PREVENT predictor range while values >3SD above the upper limit of PREVENT were ignored and deemed as likely input or coding errors.^5^ eGFR values coded as “>60” or equivalent were numerically set to 90. If an eGFR value was not recorded but a creatinine result was present, eGFR was calculated using the CKD-EPI 2021 equations.^6^

In ELSA, current smoking was defined by self-reported questionnaire answers. Diabetes Mellitus was defined as the report of a medical diagnosis, report of using hypoglycemic agents or insulin, fasting serum glucose ≥126 mg/dL, glycated hemoglobin (HbA1c) level ≥ 6.5%, or a 2-hour oral glucose tolerance test ≥ 200 mg/dL measured twice.^7^ Anthropometric and biochemical measures were recorded during baseline assessment.^4^

In the UKB current smoking was defined based on ICD10 codes or self-reported questionnaire answer. Diabetes was defined based on ICD-10 codes or an HbA1c results of >6.5%. Anthropometric and biochemical measures were recorded during baseline assessment.^2^

#### ECG preprocessing

##### Signals

Raw 12-lead ECG signals were truncated to the first 5000 samples and scaled to millivolts based on file-specific metadata information. Baseline wander was corrected using a 500-sample median filter applied independently to each lead, removing low-frequency drift. 1-lead signals were extracted by isolating lead 1 from the 12-lead signal.

##### Images

Pre-processed ECG waveforms were converted to images using a custom waveform plotting function, ECG images were plotted in standard clinical layout from signal waveform data, with a voltage calibration of 10 mm/mV, with the limbs and precordial leads arranged in 4 columns of 2.5 seconds each, representing leads I, II, and III; aVR, aVL, and aVF; V_1_, V_2_, and V_3_; and V_4_, V_5_, and V_6_. A 10-second recording of the lead I signal was included as a rhythm strip.

#### Model Architecture

##### Signal models:

We developed a 1-dimensional residual convolutional neural network (ResNet-1D) pretrained to extract features from noise-augmented ECG waveform data. The network begins with a standard “stem” block (1D convolution with a kernel size of 7, batch normalization, ReLU, and max pooling), followed by four residual stages that progressively downsample the waveform while increasing the channel depth. Each stage comprises two basic residual blocks with skip connections, facilitating stable training. After the final stage, global average pooling produces a fixed-length ECG feature vector.

##### Vision transformer model:

To extract cardiovascular representations from ECG images, our team previously utilized Contrastive Language-Image Pretraining (CLIP) to align ECG image representations with text representations from TTE reports via a dual encoder architecture.^8,9^ Up to 10 ECGs were included per patient, and they were plotted from signals in a randomized layout as a form of data augmentation. The ECG image (vision) encoder was initialized using BEiT-base-patch16-384.^10^ The TTE report (text) encoder used a CLIP text transformer.^8^ For our analysis, we extracted and finetuned the vision encoder alone.

##### Training and hyperparameter tuning:

To train our models to predict time-to-event ASCVD outcomes, we appended a multi-layer perceptron with a single output neuron to the ECG feature vector. Models weights were unfrozen and finetuned using a neural-network adapted Cox proportional hazards loss from the *TorchSurv* library, with Efron’s method for handling ties and mean reduction of the partial log-likelihood.^11–13^ The model output is then theoretically equivalent to the log-hazard for each individual, which can be converted to a survival prediction using a pre-calculated baseline survival.

The specific architecture (hidden layer dimensionality, number of hidden layers) of the multilayer perceptron, learning rate, and dropout rate were determined through hyperparameter tuning using the OPTUNA package and Weights & Biases with validation set loss in YNHHS.^14,15^ The Adam optimizer was utilized for training.^16^

### SUPPLEMENTARY TABLES

Supplementary Table 1: Specific ICD-10-CM code categories used to define ASCVD in the YNHHS and the UKB

| **Outcome** | **ICD-10-CM codes** |
| --- | --- |
| **Myocardial infarction (MI)** | **I21, I22** |
| **Hemorrhagic and Ischemic stroke** | **I61, I62, I63** |
| **Heart Failure** | **I50** |

Supplementary Table 2: Specific ICD-10-CM code categories used to define current smoking and diabetes in the YNHHS and the UKB

| **PREVENT variable** | **ICD-10-CM codes** |
| --- | --- |
| **Current smoking** | **Z72, Z87, F17** |
| **Diabetes** | **E11** |

Supplementary Table 3: Specific ATC codes used to define blood pressure-lowering and lipid-lowering medications

| **PREVENT variable** | **ICD-10-CM codes** |
| --- | --- |
| **Blood pressure-lowering medication** | **C02, C03, C07, C08, C09** |
| **Lipid-lowering medication** | **C10AA, C10B** |

Supplementary Table 4: C-index and 95% CI of different models in YNHHS and the external validation cohorts

| **Cohort** | **Model** | **C-index (95% CI)** |
| --- | --- | --- |
| YNHHS | Age + Sex (Cox model) | 0.692 [95% CI: 0.680–0.703] |
| YNHHS | ECG only model (1 lead) | 0.658 [95% CI: 0.645–0.671] |
| YNHHS | ECG only model (12 lead) | 0.692 [95% CI: 0.679–0.704] |
| YNHHS | ECG-ASCVD-1 | 0.709 [95% CI: 0.698–0.721] |
| YNHHS | ECG-ASCVD-12 | 0.711 [95% CI: 0.699–0.722] |
| YNHHS | ECG-ASCVD-IMAGE | 0.714 [95% CI: 0.703–0.726] |
| YNHHS | PREVENT | 0.712 [95% CI: 0.701–0.724] |
| YNHHS complete cases | Age + Sex (Cox model) | 0.704 [95% CI: 0.671–0.738] |
| YNHHS complete cases | ECG only model (1 lead) | 0.653 [95% CI: 0.615–0.690] |
| YNHHS complete cases | ECG only model (12 lead) | 0.696 [95% CI: 0.661–0.731] |
| YNHHS complete cases | ECG-ASCVD-1 | 0.718 [95% CI: 0.684–0.751] |
| YNHHS complete cases | ECG-ASCVD-12 | 0.723 [95% CI: 0.691–0.756] |
| YNHHS complete cases | ECG-ASCVD-IMAGE | 0.721 [95% CI: 0.688–0.755] |
| YNHHS complete cases | PREVENT | 0.724 [95% CI: 0.689–0.758] |
| ELSA | Age + Sex (Cox model) | 0.707 [95% CI: 0.665–0.748] |
| ELSA | ECG only model (1 lead) | 0.644 [95% CI: 0.597–0.692] |
| ELSA | ECG only model (12 lead) | 0.718 [95% CI: 0.674–0.763] |
| ELSA | ECG-ASCVD-1 | 0.735 [95% CI: 0.696–0.774] |
| ELSA | ECG-ASCVD-12 | 0.746 [95% CI: 0.706–0.786] |
| ELSA | ECG-ASCVD-IMAGE | 0.748 [95% CI: 0.709–0.787] |
| ELSA | PREVENT | 0.782 [95% CI: 0.746–0.817] |
| UKB | Age + Sex (Cox model) | 0.661 [95% CI: 0.642–0.681] |
| UKB | ECG only model (1 lead) | 0.592 [95% CI: 0.570–0.614] |
| UKB | ECG only model (12 lead) | 0.637 [95% CI: 0.616–0.658] |
| UKB | ECG-ASCVD-1 | 0.671 [95% CI: 0.651–0.691] |
| UKB | ECG-ASCVD-12 | 0.684 [95% CI: 0.665–0.704] |
| UKB | ECG-ASCVD-IMAGE | 0.673 [95% CI: 0.653–0.694] |
| UKB | PREVENT | 0.696 [95% CI: 0.678–0.714] |
| ASCVD indicates atherosclerotic cardiovascular disease; CI, confidence interval; ELSA, Brazilian Longitudinal Study of Adult Health (ELSA-Brasil); PREVENT, Predicting Risk of cardiovascular disease EVENTs; UKB, UK Biobank; and YNHHS, Yale New Haven Health System. | | |

Supplementary Table 5: Hazard Ratios for covariates in the PREVENT + ECG-ASCVD score models across YNHHS and the external validation cohorts

| Cohort | Model | Adjusted Hazard Ratios |
| --- | --- | --- |
| YNHHS complete case | PREVENT | 1.575 [95% CI: 1.413–1.757] |
| YNHHS complete case | ECG-ASCVD-12 | 1.338 [95% CI: 1.183–1.513] |
| ELSA | PREVENT | 1.670 [95% CI: 1.443–1.934] |
| ELSA | ECG-ASCVD-12 | 1.225 [95% CI: 1.036–1.449] |
| UKB | PREVENT | 1.432 [95% CI: 1.290–1.589] |
| UKB | ECG-ASCVD-12 | 1.331 [95% CI: 1.195–1.482] |
| ASCVD indicates atherosclerotic cardiovascular disease; CI, confidence interval; ELSA, Brazilian Longitudinal Study of Adult Health (ELSA-Brasil); PREVENT, Predicting Risk of cardiovascular disease EVENTs; UKB, UK Biobank; and YNHHS, Yale New Haven Health System. | | |

Supplementary Table 6: PREVENT variable availability in the simulated deployment cohort

| **PREVENT Variables** | **Missing (%)** |
| --- | --- |
| BMI | 56.2% |
| Total cholesterol | 81.1% |
| HDL | 81.1% |
| eGFR | 69.4% |
| BMI indicates body mass index; eGFR, estimated glomerular filtration rate; HDL, high-density lipoprotein; and PREVENT, Predicting Risk of cardiovascular disease EVENTs. | |

### SUPPLEMENTARY FIGURES

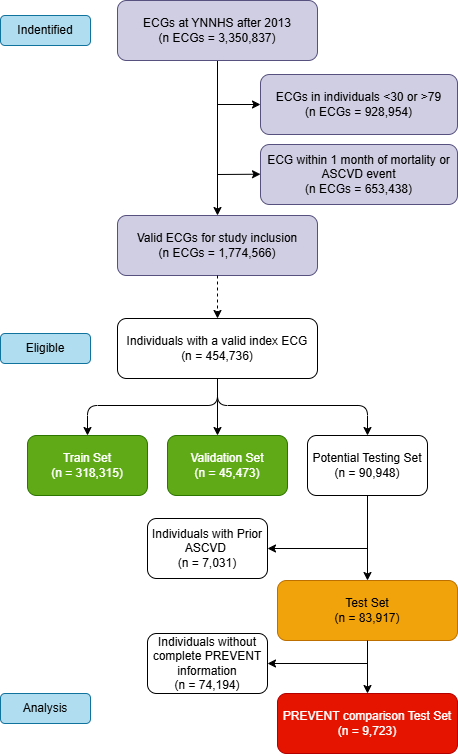

Supplementary Figure S1:Participant inclusion flowchart Yale New Haven Hospital System (YNHHS). ECG, electrocardiogram. ASCVD, Atherosclerotic cardiovascular disease. PREVENT, Predicting Risk of CVD EVENTs equations.

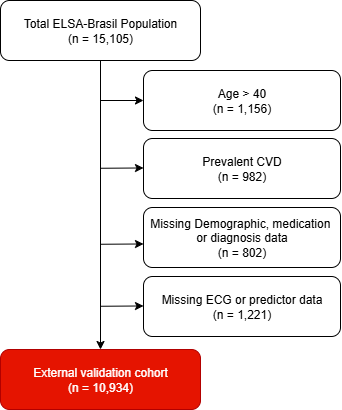

Supplementary Figure S2: Participant inclusion flowchart for the Brazilian Longitudinal Study of Adult Health (ELSA-Brasil). CVD, cardiovascular disease

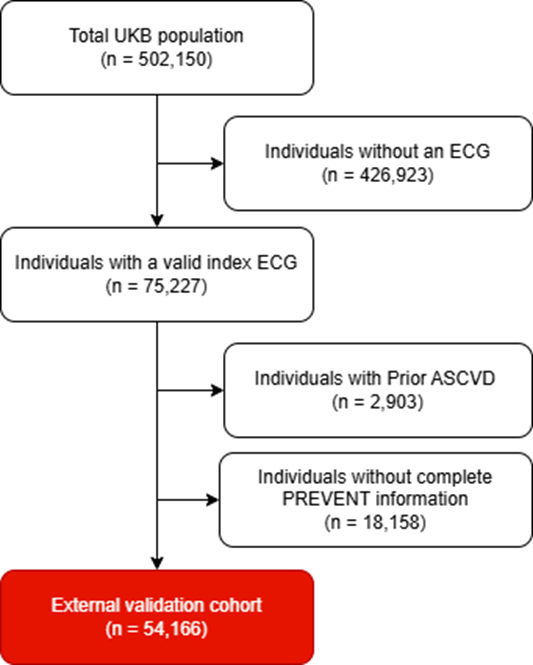

Supplementary Figure S3: Participant inclusion flowchart for the UK Biobank (UKB). ASCVD, atherosclerotic cardiovascular disease

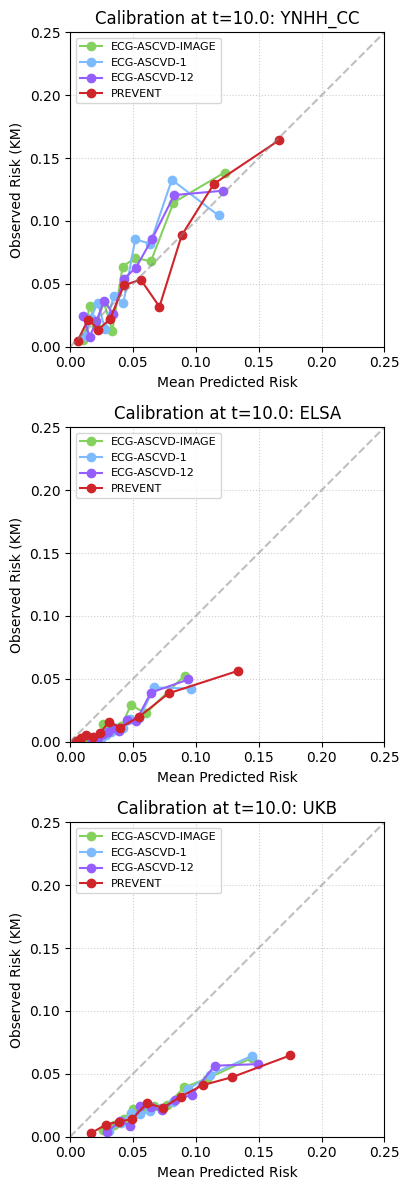

Supplementary Figure S4: ECG-ASCVD-1, ECG-ASCVD-12, ECG-ASCVD-IMAGE and PREVENT score calibration in deciles of predicted risk for predicting 10-year ASCVD rates in the YNHH complete case cohort, ELSA and UKB. Observed risk is estimated using the Kaplan Meier estimator.

**Supplement References**

1.	Palmer LJ. UK Biobank: bank on it. *The Lancet*. 2007;369(9578):1980-1982. doi:10.1016/S0140-6736(07)60924-6

2.	Littlejohns TJ, Sudlow C, Allen NE, Collins R. UK Biobank: opportunities for cardiovascular research. *Eur Heart J*. 2019;40(14):1158-1166. doi:10.1093/eurheartj/ehx254

3.	Aquino EML, Barreto SM, Bensenor IM, et al. Brazilian Longitudinal Study of Adult Health (ELSA-Brasil): objectives and design. *Am J Epidemiol*. 2012;175(4):315-324. doi:10.1093/aje/kwr294

4.	Schmidt MI, Duncan BB, Mill JG, et al. Cohort Profile: Longitudinal Study of Adult Health (ELSA-Brasil). *Int J Epidemiol*. 2015;44(1):68-75. doi:10.1093/ije/dyu027

5.	Khan SS, Matsushita K, Sang Y, et al. Development and Validation of the American Heart Association’s PREVENT Equations. *Circulation*. 2024;149(6):430-449. doi:10.1161/circulationaha.123.067626

6.	Inker LA, Eneanya ND, Coresh J, et al. New Creatinine- and Cystatin C–Based Equations to Estimate GFR without Race. *N Engl J Med*. 2021;385(19):1737-1749. doi:10.1056/NEJMoa2102953

7.	Schmidt MI, Hoffmann JF, de Fátima Sander Diniz M, et al. High prevalence of diabetes and intermediate hyperglycemia - The Brazilian Longitudinal Study of Adult Health (ELSA-Brasil). *Diabetol Metab Syndr*. 2014;6:123. doi:10.1186/1758-5996-6-123

8.	Radford A, Kim JW, Hallacy C, et al. Learning Transferable Visual Models From Natural Language Supervision. In: *Proceedings of the 38th International Conference on Machine Learning*. PMLR; 2021:8748-8763. Accessed July 17, 2025. https://proceedings.mlr.press/v139/radford21a.html

9.	Oikonomou EK, Batinica B, Dhingra LS, Aminorroaya A, Coppi A, Khera R. TARGET-AI: a foundational approach for the targeted deployment of artificial intelligence electrocardiography in the electronic health record. *medRxiv*. Published online October 25, 2025:2025.08.25.25334266. doi:10.1101/2025.08.25.25334266

10.	Bao H, Dong L, Piao S, Wei F. BEiT: BERT Pre-Training of Image Transformers. *arXiv*. Preprint posted online September 3, 2022. doi:10.48550/arXiv.2106.08254

11.	Monod M, Krusche P, Cao Q, et al. TorchSurv: A Lightweight Package for Deep Survival Analysis. *JOSS*. 2024;9(104):7341. doi:10.21105/joss.07341

12.	Cox DR. Regression Models and Life-Tables. *Journal of the Royal Statistical Society Series B: Statistical Methodology*. 1972;34(2):187-202. doi:10.1111/j.2517-6161.1972.tb00899.x

13.	Efron B. The Efficiency of Cox’s Likelihood Function for Censored Data. *Journal of the American Statistical Association*. 1977;72(359):557-565. doi:10.1080/01621459.1977.10480613

14.	Akiba T, Sano S, Yanase T, Ohta T, Koyama M. Optuna: A Next-generation Hyperparameter Optimization Framework. *arXiv*. Preprint posted online July 25, 2019. doi:10.48550/arXiv.1907.10902

15.	Biewald L. Experiment Tracking with Weights and Biases. Published online 2020. https://www.wandb.com/

16.	Kingma DP, Ba J. Adam: A Method for Stochastic Optimization. *arXiv*. Preprint posted online January 30, 2017. doi:10.48550/arXiv.1412.6980
